# ‘Drawing on Wisdom to Cope with Adversity:’ A Systematic Review Protocol of Older Adults’ Mental and Psychosocial Health During Acute Respiratory Disease Propagated-Type Epidemics and Pandemics (COVID-19, SARS-CoV, MERS, and Influenza)

**DOI:** 10.1101/2020.06.04.20122812

**Authors:** Jose M. Aravena, Cristopher Aceituno, Kate Nyhan, Kewei Shi, Sten Vermund, Becca R. Levy

## Abstract

**Background:** Mental health has become one of the fundamental priorities during the COVID-19 pandemic. Situations like physical distancing as well as being constantly tagged as the most vulnerable group could expose older adults to mental and psychosocial burdens. Nonetheless, there is little clarity about the impact of the COVID-19 pandemic or similar pandemics in the past on the mental illness, wellbeing, and psychosocial health of the older population compared to other age groups.

**Objectives:** To describe the patterns of older adults’ mental and psychosocial health related to acute respiratory disease propagated-type epidemics and pandemics and to evaluate the differences with how other age groups respond.

**Eligibility criteria:** quantitative and qualitative studies evaluating mental illness, wellbeing, or psychosocial health outcomes associated with respiratory propagated epidemics and pandemics exposure or periods (COVID-19, SARS-CoV, MERS, and Influenza) in people 65 years or older.

**Data source:** Original articles published until June 1st, 2020, in any language searched in the electronic healthcare and social sciences database: MEDLINE, Embase, CINAHL, PsycINFO, Scopus, WHO Global literature on coronavirus disease database, China National Knowledge Infrastructure (中国 知网 –CNKI). Furthermore, EPPI Centre’s COVID-19 living systematic map and the publicly available publication list of the COVID-19 living systematic review will be incorporated for preprints and recent COVID-19 publications.

**Data extraction:** Two independent reviewers will extract predefined parameters. The risk of bias will be assessed.

**Data synthesis:** Data synthesis will be performed according to study type and design, type of epidemic and pandemic, types of outcomes (mental health and psychosocial outcomes), and participant characteristics (e.g., sex, race, age, socioeconomic status, food security, presence of dependency in daily life activities independent/dependent older adults). Comparison between sex, race, and other age groups will be performed qualitatively, and quantitatively if enough data is available. The risk of bias and study heterogeneity will be reported for quantitative studies.

**Conclusion:** This study will provide information to take actions to address potential mental health difficulties during the COVID-19 pandemic in older adults and to understand responses on this age group. Furthermore, it will be useful to identify potential groups that are more vulnerable or resilient to the mental-health challenges of the current worldwide pandemic.

## Introduction

According to the World Health Organization (WHO), mental health is defined as “a state of well-being in which the individual realizes his or her own abilities, can cope with the normal stresses of life, can work productively and fruitfully, and is able to make a contribution to his or her community”.^1^ This definition considers several aspects of wellbeing and psychosocial health that are fundamental to maintain an optimal state of health. On the other hand, a relevant part of mental health acknowledges the influence of mental illness in the life of people. Mental illness is described by the American Psychiatric Association (APA) as “health conditions involving changes in emotion, thinking or behavior (or a combination of these). Mental illnesses are associated with distress and/or problems functioning in social, work, or family activities.”^2^ Under both definitions, mental health will be influenced by psychosocial situations as well as mental illness. The presence of harmful psychosocial exposures (e.g., loneliness, stigma, social isolation) and an increase in mental illness can be triggered by exposure to natural disasters that affect populational health such as epidemics and pandemics.

The recent SARS-CoV-2 virus (COVID-19) outbreak has meant a major threat to the worldwide population in several aspects of health, including mental health and psychosocial health, being an emerging significant challenge and research priority for the global population.^3^ A good point of comparison to understand COVID-19 present and future mental-health consequences are the past and present experiences observed during epidemics and pandemics outbreaks of similar characteristics. Experiences observed in other acute respiratory infections-propagated epidemics and pandemics like SARS-CoV, MERS, and influenza,^4^ have left us a precedent of information regarding its substantial impact on people’s mental health. Situations such as physical distance as one of the most critical measures, uncontrolled exposure to media news about the virus, spread biased or false information, quarantine, isolation, economic hardships, loss of love ones, health consequences, burden, stigma, fear, and anxiety; consequences that have been observed in the present and passed epidemic and pandemic scenarios.^5–8^ Therefore, these experiences must be carefully considered to generate an early response at an individual and populational level, and to anticipate prospective mental health scenarios. In that regard, recently Rogers and Cols have observed through a systematic review and meta-analysis of psychiatric and neuropsychiatric consequences associated with coronaviruses infections^9^ that among patients with severe SARS or MERS coronavirus infections, delirium, post-traumatic stress disorder, depression, anxiety, and fatigue are common. Moreover, in some preliminary data, COVID-19 would present delirium as well as confusion, agitation, depressive symptoms, anxiety, and insomnia. This study set an important precedent about how impactful the coronavirus infection in mental health could be. Although, the study did not include the contextual impact of epidemic and pandemics, the full range of psychosocial and wellbeing aspects, and did not compare the mental health among different ages. Areas that must be analyzed to understand the full range of influences in mental health and experiences across age groups.

A group that could be highly affected are those who have been categorized as high-risk to present severe symptoms or mortality related to the virus such as people with chronic diseases and groups of older adults. COVID-19 pandemic has demonstrated to be a critical challenge for older people’s physical health. People 65 years or older are the population with the highest risk of mortality associated with COVID-19 worldwide.^10^ Patients with multimorbidity and cardiovascular risk, which increase exponentially after 65 years old, are particularly prone to manifest severe symptoms.^11–13^ Thus, many communities have suggested or enforced particularly strict prevention measures for older persons with these characteristics.

Mental health burden could be an associated consequence of being the population at the highest risk and the exposure to strict social isolation in a pandemic. COVID-19 virus and its preventive methods imply important mental health challenges for older people and caregiver’s health that must be addressed on time. The classification of “population of high-risk” or in need of shielding could be a source of stress and stigma for older adults, incrementing its social isolation and mental illness symptoms such as anxiety or depression.^14,15^ Mental health burden is particularly harmful to older adults with some degree of dependence in daily life activities or multimorbidity because they manifest a higher risk to experience increased physical frailty and worsening of other diseases.^16–20^ If mental illness symptoms and psychosocial difficulties increase in the frail and geriatric older adult’ populations during a pandemic period, the rise of dependency, chronic diseases, and emergency visits for causes other than COVID-19 would be an enormous collateral impact of the current worldwide pandemic.

Diverse and often underlooked realities of aging constitute older adulthood, from independent older adults who have not stopped their work activity, caregivers of family members (e.g., other older adults, grandchildren), older people living on their own, or heads of household, to older persons who require the support of a third person, or others who live in long-term care institutions. In this context, older adults’ mental health during natural disasters is controversial. Some studies about resilience in other contexts have shown that older adults tend to report a higher resilience and more positive outcome than other age groups,^21,22^ and others have estimated that older adults are 2.11 and 1.73 more likely to experience PTSD and adjustment disorder symptoms after natural disasters compared to younger adults, respectively.^23^ Nevertheless, under normal circumstances, the evidence has shown that older people then to manifest greater levels of wellbeing, lower levels of negative affects, and less distress during their social interactions than other age groups.^24^ Furthermore, studies have evidenced that older adults are more prone to put attention to positive stimulus than negative ones compared to younger people that present opposite patterns, putting more focus on negative situations.^25,26^ This talks about certain ability to allocate emotional resources that could be fundamental to cope in a more positive manner with unpredictable or emotionally demanding events.^27^

Despite all of these, there has not yet been a systematic evaluation to understand these patterns in the context of epidemics or pandemics. Therefore, although older adults have been constantly classified as a vulnerable population for COVID-19, there exists uncertainty about how older adults, compared to other age groups, could respond to a situation that requires an important mental endurance like an epidemic or pandemic.

Published and ongoing studies, such as Roger et al,^9^ who have characterized the mental illness and neuropsychiatric consequences associated to coronavirus infections in the general population, and Qin and cols who have registered a protocol for a meta-analysis of the impact of COVID-19 on the mental wellbeing of elderly population,^28^ have focused their reviews just on clinical outcomes related to mental health. In this context, and considering the increasing number of COVID-19 related articles, a systematic review targeted to older people mental health considering a full-range of neuropsychiatric, psychiatric, psychosocial, and wellbeing parameters associated with the infection or the contextual impacts related to acute respiratory disease propagated-type epidemics and pandemics, contrasting the results among groups seems pertinent and necessary to fully understand the response and experiences of older adults and other age groups in the context of pandemics.

To comprehend what could be the potential mental health impact associated with respiratory propagated epidemics and pandemics in older adults, and to evaluate the contrast among different age groups it is critical information for the development and planning of policies and programs to address these consequences early and to understand intergenerational differences and similarities in the mental health response to epidemic and pandemics. At the same time, it is fundamental information for the development of interventions and the implementation of policies targeted to change or promote behaviors related to compliance of nonpharmacological measures to prevent the spread of acute respiratory diseases during the context of epidemics and pandemics.

### Study aims

Considering this background, the main goal of this review is to describe the patterns of older adults’ mental health related to acute respiratory disease propagated-type epidemics and pandemics.

Specifically, this systematic review aims 1) to describe the associations between respiratory propagated epidemic and pandemics and older adult’s mental health, 2) to describe the differences between older adults and other age groups in the effects of mental health factors related to acute respiratory disease propagated-type epidemics and pandemics periods in the mental health, 3) to assess the effect of interventions in the older adult’s mental health associated to respiratory propagated epidemic and pandemics, and 4) to consider moderators of the impact of pandemics on older adults’ mental health.

## Method

### Type of studies

The report of the study will follow the PRISMA statement for reporting systematic reviews and meta-analyses guidelines.^29^ We will select studies that: 1) describe the effects of acute respiratory disease propagated-type epidemics or pandemics on mental health or psychosocial parameters, and 2) include older adults in the sample. Quantitative, qualitative, and mixed-method studies will be included in order to consider different aspects of mental health and psychosocial impact.

### Type of participants

Any study evaluating people 60 years or older residing in any setting. Research involving people from other age groups (e.g. children, adolescents, adults) additionally to people 60 years or older will be included for analysis.

### Types of exposure

For this review, studies conducted evaluating the impact on mental health during defined acute respiratory disease propagated-type epidemic or pandemic according to the *Infection prevention and control of epidemic-and pandemic prone acute respiratory infections in health care: WHO guidelines. 2014*:^4^ SARS coronavirus (SARS-CoV), Middle East Respiratory Syndrome (MERS), and Influenza/flu (H1N1, H5N1). SARS coronavirus 19 (SARS-CoV-2 or COVID-19) will be also included. These viruses are selected because they share similar epidemiological characteristics, where its pathogens can cause large scale outbreaks with high morbidity and mortality.^4^

### Types of outcomes measures

For the purpose of this review, any study describing outcomes associated with mental health parameters in older adults will be included. Mental health will be understood under the WHO definition: “a state of well-being in which the individual realizes his or her own abilities, can cope with the normal stresses of life, can work productively and fruitfully, and is able to make a contribution to his or her community.”^1^ For practical operationalization, it will be divided into two main components: mental illness and psychosocial health/wellbeing. Examples of mental illness parameters are depression, anxiety, and mood disorders, including intervention studies. Studies analyzing parameters such as cognition, dementia, and delirium would be incorporated under the umbrella of mental illness aspects because people with these diagnoses frequently manifest neuropsychiatric symptoms. Examples of psychosocial health/wellbeing factors are quality of life, stigma, isolation, and loneliness. Studies evaluating the mental illness and psychosocial health/wellbeing parameters of caregivers of older adults will be incorporated.

### Search method for identification of studies

Original articles published until June 1st, 2020, in any language searched in the electronic healthcare and social sciences databases: MEDLINE (Ovid), Embase (Ovid), CINAHL (Ebsco), PsycINFO (Ovid), Scopus, WHO Global literature on coronavirus disease database, China National Knowledge Infrastructure (中国知网 – CNKI). Because of limitations in database coverage and indexing speed, COVID-19 related articles will be identified in two other ways. First, studies in the EPPI Centre COVID-19 living systematic map of the evidence screening review^30^ which are tagged with “health impacts,” “social/economic impact,” or “mental health impacts” will be added to the screening workflow. The EPPI Centre COVID-19 map consists of studies on COVID-19, identified in MEDLINE and Embase, and published in 2019 or later. Second, for better coverage of preprints, we will use the publicly available publication list of the COVID-19 living systematic review^31^, which retrieves articles from the preprints databases bioRxiv and medRxiv and it is continuously updated.

Because more COVID-19 related articles are published week by week, after the title-abstract screening is completed, another search exclusively for COVID-19 related-articles will be performed in order to include manuscripts that potentially were published or indexed after the date of the first round of database searches. Articles included from this second COVID-19 related-articles extraction will be screened in the same fashion as the other studies.

An example of the MEDLINE search strategy and a search source scheme are described in the supplement section. The search will be adjusted for appropriate controlled vocabulary and syntax in each database. In each database, the search has three elements: queries for the exposure of interest (COVID-19 or other respiratory-propagated pandemics), the outcomes of interest (mental health), and the population of interest (older adults). Controlled vocabulary and indexing status will be used, where possible, to maximize the retrieval of papers dealing with the older adult population and to minimize the burden of screening papers about other age groups.

No specifications about the type of study are included in the search strategy to reduce the risk of missing studies. Mental illness terms were included following the DSM-V and the Cochrane Common Mental Disorders group search strategies (https://cmd.cochrane.org/). Some psychosocial health/wellbeing terms were incorporated from other systematic reviews about psychosocial health and wellbeing and based on expert opinion.^32,33^ Because an important part of the epidemics and pandemics of these viruses has been experienced in the Chinese population, culturally sensible terms to describe mental illness (‘Impulsive personality disorder,’ ‘Qigong-induced disorders,’ ‘Traveling psychosis,’ ‘*Shenjing shuairuo*,’ and ‘Neurasthenia’) and psychosocial health/wellbeing conditions (‘Shame,’ ‘Humiliation,’ ‘Low spirits,’ ‘Witchcraft,’ ‘Curses,’ ‘*Zou huo ru mo* –走火入魔- or Qigong deviation –氣功偏差-‘) were included.^34,35^

Studies will be divided into two main categories for its analysis: 1) studies describing the direct effect of virus infection on mental health outcomes, and 2) studies illustrating mental health impact associated with the contextual situation of the epidemic or pandemic (e.g. quarantines, social distancing, isolation).

### Data collection and analysis

The results from all the database searches will be collated in EndNote and deduplicated by the Cushing/Whitney Medical Library Cross-Departmental Team. The deduplicated results will be uploaded to Covidence, an online platform for evidence synthesis. Reviewers (JA and CA) will screen articles at the title abstract level, discarding only those articles which are evidently off-target. The full-text screening will also take place in Covidence. Two independent screeners will vote on each article; disagreements will be solved by consensus or third-party adjudication (BL). Articles in English and Spanish language will be manipulated by two reviewers (JA and CA). Articles in other languages will be handled by two research members (KS and SV).

### Data extraction and management

Two independent reviewers will perform data extraction using a prespecified data abstraction form designed for this study. The data abstraction form will be pilot-tested on five randomly-selected studies and refined accordingly. Data extraction will include characteristics of the study (e.g. country, data source, data collection date, year), methods (e.g. study design, sample characteristics, outcome measurement), and results. Extracted studies will be tagged according to the type of outcome they are describing: a) virus infection mental health-related outcomes, b) epidemic or pandemic context mental health-related outcomes, or c) both types of outcomes. Data will be entered in a duplicated Google questionnaire specifically designed for the study. Every researcher will enter the data on independent questionnaires.

### Assessment of risk of bias in included studies

Qualitative and mixed-method studies will be described. Quantitative studies will be included for assessment of the risk of bias. Two reviewers will independently assess the internal validity of each included quantitative study. Study risk of bias will be categorized as low risk of bias, some concerns of bias, and high risk of bias.

In the case of observational studies, bias will be evaluated following the next standards: 1) Ttype of study design, 2) Temporality of the evaluation of the exposure: concordance in the evaluation timing of the impact of the epidemic/pandemic episode with the study goals, 3) Outcome evaluation: evaluation of the outcome with standardized and defined measurement instrument or methods, 4) Adjusted analysis: the inclusion of an adjusted analysis of the main outcome considering relevant variables. For this review, analyses adjusting for age, sex, and pre-existing medical conditions or functional performance will be considered acceptable. 5) Attrition bias: for cohort studies, 30% of loss of follow-up will be considered as acceptable.

For intervention studies evaluating efficacy or effectiveness in one or more mental health and psychosocial health as a primary outcome, the criteria to evaluate the risk of bias will be: 1) Type of study design, 2) Bias arising from the randomization process, 3) Bias due to deviations from intended interventions, 4) Bias due to missing outcome data, 5) Bias in measurement of the outcome, and 6) Bias in selection of the reported result. Studies incorporating mental health parameters as secondary outcomes will be included for description yet will be considered at a high risk of bias.

Observational study’s risk of bias was designed considering STROBE and the AHRQ Methods guidelines.^36,37^ Intervention study risk of bias follows the Cochrane Handbook for Systematic Reviews.^38^

### Measures of treatment effect

In the case of quantitative studies, for the continuous variables related to mental health, because of the variety of scores and outcomes produced by the diverse measurement scales, measures such as frequency and prevalence of symptoms and diagnosis (%) or adjusted prevalence, mean and standard deviation (SD) of total scores will be used. In comparison studies, mean differences (MD), proportions (%), standardized mean differences (SMD), B coefficient, and standardized error, with 95% confidence intervals (CI) for continuous outcomes will be included. Dichotomous outcomes such as adjusted risk ratios (RR), Odds ratio (OR), and Hazard ratio (HR) with 95% CIs will be considered. Unadjusted and adjusted results will be extracted. These measures will be extracted for people 65 years older, other age groups described in every article, sex, and race if it is included.

### Unit of analysis issues

For treatment, in the case of cluster randomized trials or interventions delivered in groups, the unit of analysis will be the cluster. For interventions including individuals, the unit of analysis will be the subjects.

### Dealing with missing data

In the case of RCTs, we will seek data irrespective of compliance, in order to allow the intention to treat analysis. For cohort studies, we will make a qualitative evaluation of every study to identify if the missed data lead to a bias in the result.

### Assessment of heterogeneity

We will judge heterogeneity among studies (the type of study design, inclusion criteria, type of exposure/intervention, outcome measurement) during the qualitative synthesis of the data. Additionally, statistical heterogeneity was evaluated using the I2 statistics, classifying no heterogeneity (< 25%), low (25–49%), moderate (50–74%), and high heterogeneity (equal or > 75%). We will decide on the appropriateness of conducting a meta-analysis based on qualitative and quantitative information.

### Assessment of reporting bias

To avoid publication bias, we will search for published studies in multiple databases which include published journal articles and preprints. Every study will be evaluated and discussed considering its bias and strengths for inclusion in the review. We will report the number of articles that do not fulfill requirements. For studies with two documents (preprint and journal publication), the official publication will be considered. In the studies with more than one analysis, the most tailored to our study aim publication will be considered. Funnel plots will be performed for publication bias if we have enough data.

### Data synthesis and results

A descriptive analysis of the included studies will be conducted through a flow diagram describing the number of included and excluded studies, exclusion reasons (e.g. older population not included, different epidemic/pandemic exposure, non-mental health outcomes), and the final number of selected studies. The results will be synthesized in tables and figures which may include the following. Table 1 will display study characteristics (country, data source, data collection dates, year, type of study/study design, total sample by group, follow-up, participants basic characteristics, exposed epidemic/pandemic), table 2 outcome measurement (name of the outcomes, type of outcome –mental health/psychosocial-, outcome measurement, and results). A third table will describe intervention studies and its results (country, data collection and intervention delivery dates, year, type of research design, inclusion/exclusion criteria, description of the intervention, exposed epidemic/pandemic, sample by group, intervention/control characteristics, outcome measurement, results). Data synthesis will be performed according to study type and design, type of epidemic and pandemic, types of outcomes (mental health and psychosocial outcomes), and participant characteristics (sex, race, comparison to other age groups, independent/dependent older adults). Comparison between sex, race, and other age groups will be performed qualitatively, and quantitatively if the data available is enough. The risk of bias and heterogeneity will be reported for quantitative studies published in journal articles or preprints.

### Subgroup analysis and investigation of heterogeneity

If the available data is enough, we plan to conduct a subgroup analysis considering the following categories: type of study design, type of outcome measured, type of epidemic, or pandemic. If the data available is enough quantitative comparison of age groups will be conducted.

### Sensitivity analysis

We will perform a sensitivity analysis based on studies with a low risk of bias.

## Discussion

Mental health understood as a state of wellbeing has been a topic of special discussion and concern in the health and medical sciences because of its impact on the people’s lives and the high burden for societies. In the context of large-scale natural disasters such as epidemics and pandemics, mental health would be highly determined by the manifestation of mental illnesses, neuropsychiatric conditions, and psychosocial aspects that will influence people’s health and their capacity to cope with a mentally demanding situation.^3^ This topic takes major relevance in the current scenario triggered by the COVID-19 worldwide pandemic, where there exist and evident relevance of understanding the patterns of mental coping and adaptation of the global population.

In our actual society, people 65 years or older have been increasingly exposed to situations that are a threat to their mental health such as isolation and loneliness.^39^ At the same time, the constant exposure to ‘ageism’ or negative stereotypes associated with the aging as well as classifications of ‘population of high-risk’ or in need of shielding could be an important source of stress, fear, and segregation. Nevertheless, even in the presence of these negative ideas about older people, the evidence has been uncertain about older adult’s mental resilience and adaptation compared to other age groups in front of natural disasters. Under normal situations, older adults have shown that they report higher general wellbeing and satisfaction with social connection than the younger groups.^24^

To our knowledge, this is the first systematic review evaluating the older adult’s mental and psychosocial health compared to other age groups in the context of acute respiratory disease epidemics and pandemics. Therefore, to understand how mental and psychosocial health could change during epidemics and pandemics of similar characteristics than COVID-19 in older adults in contrast to other ages will be critical to elucidate the natural emergence of mental and behavioral coping mechanisms across life-stages, and to comprehend the major necessities referred by these groups. This information will be critical for the design of interventions and policies oriented to increment positive behavioral changes across age population groups and to promote the adherence to nonpharmacological preventive measures during epidemics and pandemics.

## Data Availability

The study is a protocol of a systematic review that will use publicly available data.

